# Mid-Regional pro-Adrenomedullin (MR-proADM), C-Reactive Protein (CRP) and Other Biomarkers in the Early Identification of Disease Progression in COVID-19 Patients in the Acute NHS Setting

**DOI:** 10.1101/2021.04.19.21252978

**Authors:** Nathan Moore, Rebecca Williams, Matilde Mori, Beatrice Bertolusso, Gabrielle Vernet, Jessica Lynch, Peter Philipson, Thomas Ledgerwood, Stephen P. Kidd, Claire Thomas, Veronica Garcia-Arias, Michelle Young, Kordo Saeed, Kirsty Gordon, Nicholas Cortes

## Abstract

**Aims:** There is a lack of biomarkers validated for assessing clinical deterioration in COVID-19 patients upon presentation to secondary or tertiary care. This evaluation looked at the potential clinical application of C-Reactive Protein, Procalcitonin, Mid-Regional pro-adrenomedullin (MR-proADM) and White Cell Count to support prediction of clinical outcomes.

**Methods:** 135 patients presenting to Hampshire Hospitals NHS Foundation Trust between April and June 2020 confirmed to have COVID-19 via RT-qPCR were included. Biomarkers from within 24 hours of admission were used to predict disease progression by Cox regression and area under the receiver operating characteristic (AUROC) curves. The endpoints assessed were 30-day all-cause mortality, intubation and ventilation, critical care admission and non-invasive ventilation (NIV) use.

**Results:** Elevated MR-proADM was shown to have the greatest ability to predict 30-day mortality adjusting for age, cardiovascular, renal and neurological disease. A significant association was also noted between raised MR-proADM and CRP concentrations and the requirement for critical care admission and non-invasive ventilation.

**Conclusions:** The measurement of MR-proADM and CRP in patients with confirmed COVID-19 infection upon admission shows significant potential to support clinicians in identifying those at increased risk of disease progression and need for higher level care, subsequently enabling prompt escalation in clinical interventions.

## Introduction

Since initial reports of a cluster of pneumonia cases arising from Wuhan, Hubei and the subsequent identification of the causative *betacoronavirus*, the severe acute respiratory syndrome coronavirus 2 (SARS-CoV-2) has infected over 100 million people globally and directly resulted in the deaths of approximately 2·3 million by January 2021.

The progressive multi-organ failure associated with SARS-CoV-2 mortality is driven in part by significant inflammation and microvascular thrombosis. Presence of SARS-CoV-2 within the endothelium can cause a secondary endotheliitis and an impairment of vascular blood flow, a pro-thrombotic state and vascular leakage.^1^ During endotheliitis there is elevation of biomarkers, such as neutrophil extracellular traps, that can lead to microvascular thrombosis and inflammation.^2^

Adrenomedullin is a peptide that has been shown to have a role in preserving the integrity and stability of the endothelium after severe infection. Furthermore, it has potent vasodilatory properties, as well as having a role in immunomodulation and metabolic regulation. It is upregulated in several pathogenic processes, including sepsis and the progression of septic patients towards multi-organ failure.^3-5^

The novel biomarkers mid-regional proadrenomedullin (MR-proADM) has also been shown to be predictive of poor clinical outcomes in patients with sepsis and infections involving the respiratory^6,7^ and urinary^8,9^ systems. Similarly, it has been demonstrated during non-infective processes such as heart failure and kidney injury.^10^ As a precursor amino acid sequence that splits from proadrenomedullin, MR-proADM is used as a surrogate marker for adrenomedullin. It can be measured via an automated immunofluorescent assay, as levels are directly proportional to adrenomedullin which rapidly breaks down in blood.

The instigation of an appropriate management plan is dependent on an adequate assessment of the infection severity, which is determined by the host pathophysiological response and the virulence of the organism causing infection. However, infection is a dynamic process with infection severity changing over time within an individual. As infection progresses, patients may transition from low severity to high severity which if not identified, may lead to delayed or inappropriate therapy. Therefore, biomarkers that could help stratify risk of disease progression are of clinical interest as they may help guide appropriate early management and patient placement within hospital. A recent study by Saeed *et al*. found that the measurement of MR-proADM could effectively predict disease progression; identifying those at increased risk of mortality, those who would need ICU admission and those requiring a longer length of stay in hospital. Furthermore, they identified its potential use in facilitating early discharge from hospital, with no increase in mortality.^11^

The aim of this study was to assess the effectiveness of a number of biomarkers, both established and emerging, in the acute setting as prognostic markers to support clinicians in the early identification of patients with Covid-19 at risk of mortality, ICU admission and ventilation.

## Methods

### Study Design and Data Collection

In this observational study, we identified patients confirmed to have SARS-CoV-2 at a district general hospital, Hampshire Hospitals NHS Foundation Trust, during the first wave of the virus between April and June 2020. SARS-CoV-2 was detected by real-time reverse-transcription PCR (rRT-PCR) using the COVID-19 genesig® Real-Time PCR assay (Primerdesign Ltd, Chandler’s Ford, UK). This resulted in 135 eligible consecutive patients. All blood samples analysed were collected as part of routine clinical care on admission to hospital. Measurement of C-reactive protein (CRP) on serum samples was performed on ADVIA 2400 analysers; procalcitonin (PCT) on Centaur XP analysers and full blood count (FBC) and differential white blood cell counts were performed on Ethylenediaminetetraacetic acid (EDTA) samples on Siemens Advia 2120i (all Siemens Healthineers, Tarrytown, New York, USA). Measurement of MR-proADM was performed (within 72hours of collection, in line with manufacturer’s guidance) on EDTA blood samples taken on admission using an immunoassay (B.R.A.H.M.S. KRYPTOR™, Thermo Fisher Scientific, Henningsdorf, Germany). Those performing the assays were blinded to clinical outcomes. Where these variables had not been requested by the responsible clinician attempts were made to locate the original sample and perform the assay, within twenty-four hours.

Patients were followed up until discharge, with demographics, comorbidities, admission and discharge dates, non-invasive ventilation (NIV) use, need for intubation and ventilation, admission to ICU and mortality data being anonymously collected. Clinicians caring for the patients were unaware that outcome data was being collated in relation to the above biomarkers.

### Statistical Analysis

Cases and controls were determined by outcome variables during univariate and multivariate Cox Regression. However, this was only performed for clinical outcomes with a sufficiently high event per variable ratio.^12,13^ Time to event analysis was right-censored at 30 days. The Cox proportional hazard models were checked for proportionality by correlating sets of scaled Schoenfeld residuals with time, to test for independence between residuals and time. Where proportionality was not met stratification was performed as necessary. Hazard ratios and C-index score were calculated with corresponding 95% confidence intervals. Confounding variables for the multivariate analysis were selected based on those that were significant upon univariate logistic regression following Bonferroni correction, with a p-value of <0·005.

Symmetrically distributed variables were reported using the mean (standard deviation), whereas variables exhibiting a skewed distribution were reported using the median [first quartile – third quartile]. The Chi-square test (χ2) was used to assess differences in categorical variables in the demographics and clinical characteristics between those who did and did not die within 30 days, with the Mann-Whitney U test being used to assess differences in continuous variables and Student’s T-test for age.

Data analysis was performed using R version 4·0.1.

Receiver operating characteristic (ROC) curves with area under the curve (AUC) analysis was used to determine predictive value for each clinical endpoint with 95% confidence intervals to establish significance. To investigate if individual biomarkers offer a significant improvement in predicting an adverse outcome over a model incorporating previously identified confounding variables, as outlined above, AUC analyses were compared using a bootstrap method.

## Results

Samples from 135 patients in total were analysed with a mean age of 64·6 years and a predominance of males (51·9% male vs 48·1% female). The patient group included 105 who had survived by thirty days post admission and 30 who had died by this timepoint. The characteristics of the patient cohort are described in Supplementary Table 1.

Median biomarker concentrations in this group showed elevated median CRP 59mg/L (Q1-Q3 range 17 – 143) and MR-proADM 1·02 nmol/L (Q1-Q3 range 0·71 - 1·61); median levels within normal clinical ranges for total white blood cell count 7·1×10^9^/L (Q1-Q3 range 5·2 – 9·65), neutrophils 5·31 ×10^9^/L (Q1-Q3 range 3·51 – 7·66) and PCT 0·13ng/mL (Q1-Q3 range 0·07 - 0·41) but a suppressed median lymphocyte count 0·98 ×10^9^/L (Q1-Q3 range 0·70 – 1·36).

Twenty-five of these patients were enrolled into the RECOVERY trial with 9 receiving standard care, 6 received hydroxychloroquine, 6 received Lopinavir-Ritonavir and 3 received Azithromycin, all of which were deemed to have no clinical benefit.^14-16^ One patient received dexamethasone as part of the RECOVERY trial. Otherwise, none of the patients received other immunomodulation, including IL-6 inhibitors (tocilizumab or sarilumab), IL-1 inhibitors (such as anankinra) or convalescent plasma. Multivariate Cox regression adjusted for age, cardiovascular disease, renal disease and neurological disease. Proportionality was checked for all Cox regression models and stratification performed as necessary.

### MR-proADM

For this novel biomarker Saeed et al^11^ have previously derived and proposed, in a non-COVID-19 acute patient population, a cut-off of 1·54 nmol/L as a predictor of elevated mortality, higher rate of ICU admission and longer length of stay. Applying this cut-off to our COVID-19 patient population shows significantly (p = 1·59×10^−5^) lower mortality rate of 11·11% (N=11) in the patients with MR-proADM values < 1·54 nmol/L (N=99 (73·33%)) compared to a higher mortality rate of 52·78% (N=19) in patients (N=36, (26·67%)) with MR-proADM values equal to or exceeding a cut-off of 1·54 nmol/L. None of the patients who died (N=30) had a MR-proADM value below the pre-defined threshold of < 0·88 nmol/L (p = 1·61 × 10^−9^), previously proposed in a non-COVID-19 population by Saeed et al^11^ as a potential cut-off for identifying patients at very low risk for deterioration and no mortality.

Of interest, fifty-two (38·52%) patients with MR-proADM values of <0·88 nmol/L had a median inpatient stay of 4 days (interquartile range [IQR] 0·75 – 9 days), representing a significantly shorter length of stay than those with an MR-proADM of >0·88 nmol/L whose median stay was 12 days (IQR 5 – 22·5 days), p <0·00001.

### 30 Day Mortality

Supplementary Table 1 compares the patient demographics and clinical characteristics between survivors and non-survivors. Of note CRP, PCT and MR-proADM were all significantly raised amongst those that did not survive at day 30.

Univariate Cox regression analysis found MR-proADM to have the greatest ability to predict 30 day mortality (Wald 20·3, HR 1·5455 [1·279-1·868]), with CRP also proving effective at predicting 30 day mortality (Wald 4·02, HR 1·0034 [1·000-1·007]) as shown in Table 2.

**Table 1.** Univariate Cox regression for prediction of 30-day mortality, requirement of critical care, NIV and intubation and ventilation using biomarkers in baseline bloods

| Biomarker | Patients (N) | Cases (N) | Wald | D.F. | C-index [95% CI] | HR [95% CI] | p-value |
| --- | --- | --- | --- | --- | --- | --- | --- |
| **Prediction of 30-day mortality** |  |  |  |  |  |  |  |
| MR-proADM | 135 | 30 | 20.3 | 1 | 0.811 [0.746-0.876] | 1.5455 [1.279-1.868] | 6.64e-6 |
| CRP<sup>1</sup> | 135 | 30 | 4.02 | 1 | 0.613 [0.515-0.711] | 1.0034 [1.000-1.007] | 0.0449 |
| PCT<sup>2</sup> | 116 | 26 | 11.54 | 1 | 0.621 [0.511-0.731] | 1.0898 [1.037-1.145] | 0.000681 |
| Neutrophils | 135 | 30 | 0.73 | 1 | 0.549 [0.445-0.653] | 0.9555 [0.861-1.061] | 0.394 |
| Lymphocytes | 135 | 30 | 3.53 | 1 | 0.448 [0.338-0.558] | 1.0920 [0.996-1.197] | 0.0603 |
| **Prediction of requiring critical care admission** |  |  |  |  |  |  |  |
| MR-proADM | 135 | 31 | 3.2 | 1 | 0.614 [0.518-0.710] | 1.2570 [0.978-1.615] | 0.0737 |
| CRP<sup>1</sup> | 135 | 31 | 15.83 | 1 | 0.716 [0.624-0.808] | 1.0055 [1.003-1.008] | 0.0000695 |
| PCT<sup>2</sup> | 116 | 31 | 0.03 | 1 | 0.687 [0.595-0.779] | 1.0050 [0.951-1.062] | 0.86 |
| Neutrophils | 135 | 31 | 0.37 | 1 | 0.609 [0.511-0.707] | 1.0183 [0.960-1.080] | 0.544 |
| Lymphocytes | 135 | 31 | 2.52 | 1 | 0.465 [0.357-0.573] | 1.0759 [0.983-1.178] | 0.113 |
| **Prediction of requiring NIV** |  |  |  |  |  |  |  |
| MR-proADM | 135 | 23 | 11.62 | 1 | 0.726 [0.634-0.818] | 1.4992 [1.188-1.892] | 0.000651 |
| CRP<sup>1</sup> | 135 | 23 | 15.53 | 1 | 0.725 [0.617-0.833] | 1.0061 [1.003-1.009] | 0.0000813 |
| PCT<sup>2</sup> | 116 | 22 | 0.1 | 1 | 0.772 [0.682-0.862] | 1.0098 [0.952-1.072] | 0.747 |
| Neutrophils | 135 | 23 | 0.06 | 1 | 0.432 [0.324-0.540] | 0.9883 [0.901-1.085] | 0.804 |
| Lymphocytes | 135 | 23 | 4.81 | 1 | 0.552 [0.442-0.662] | 1.0991 [1.010-1.196] | 0.0283 |
| **Prediction of intubation and ventilation** |  |  |  |  |  |  |  |
| MR-proADM | 135 | 16 | 12.37 | 1 | 0.748 [0.650-0.846] | 1.6015 [1.232-2.082] | 0.000435 |
| CRP<sup>1</sup> | 135 | 16 | 16.57 | 1 | 0.745 [0.614-0.876] | 1.0069 [1.004-1.010] | 0.0000469 |
| PCT<sup>2</sup> | 116 | 16 | 0.63 | 1 | 0.836 [0.746-0.926] | 1.0215 [0.969-1.077] | 0.429 |
| Neutrophils | 135 | 16 | 0.43 | 1 | 0.641 [0.514-0.768] | 1.0259 [0.950-1.107] | 0.513 |
| Lymphocytes | 135 | 16 | 0.09 | 1 | 0.468 [0.337-0.599] | 0.9415 [0.630-1.408] | 0.769 |

**Table 2.** Multivariate Cox regression for prediction of 30-day mortality, requirement of critical care, NIV and intubation and ventilation using biomarkers in baseline bloods

| Biomarker | Patients (N) | Cases (N) | Wald | D.F. | C-index [95% CI] | HR [95% CI] | p-value |
| --- | --- | --- | --- | --- | --- | --- | --- |
| **Prediction of 30-day mortality** |  |  |  |  |  |  |  |
| MR-proADM | 135 | 30 | 23.736 | 5 | 0.827 [0.756-0.898] | 1.8709 [1.454-2.407] | 1.1e-6 |
| CRP<sup>1</sup> | 135 | 30 | 12.166 | 5 | 0.788 [0.704-0.872] | 1.0069 [1.003-1.011] | 0.000487 |
| PCT | 116 | 26 | - | - | - | - | - |
| Neutrophils | 135 | 30 | 0.9235 | 5 | 0.808 [0.745-0.871] | 0.9445 [0.841-1.061] | 0.336303 |
| Lymphocytes | 135 | 30 | 6.6564 | 5 | 0.791 [0.713-0.869] | 1.1213 [1.028-1.223] | 0.009879 |
| **Prediction of requiring critical care admission** |  |  |  |  |  |  |  |
| MR-proADM | 135 | 31 | 10.151 | 5 | 0.713 [0.621-0.805] | 1.5451 [1.182-2.019] | 0.00144 |
| CRP<sup>1</sup> | 135 | 31 | 22.062 | 4 | 0.775 [0.702-0.848] | 1.0078 [1.005-1.011] | 2.64e-6 |
| PCT<sup>2</sup> | 116 | 31 | 0.6084 | 5 | 0.715 [0.631-0.799] | 1.0254 [0.963-1.092] | 0.43564 |
| Neutrophils | 135 |  | 0.5761 | 5 | 0.686 [0.596-0.776] | 1.0221 [0.966-1.081] | 0.44772 |
| Lymphocytes | 135 | 31 | 1.4066 | 5 | 0.689 [0.599-0.779] | 1.0638 [0.961-1.178] | 0.23544 |
| **Prediction of requiring NIV** |  |  |  |  |  |  |  |
| MR-proADM | 135 | 23 | 30.393 | 5 | 0.828 [0.740-0.916] | 2.5230 [1.816-3.506] | 3.53e-8 |
| CRP<sup>1</sup> | 135 | 23 | 18.293 | 5 | 0.799 [0.725-0.873] | 1.0081 [1.004-1.012] | 1.89e-5 |
| PCT<sup>2</sup> | 116 | 22 | 1.39 | 5 | 0.755 [0.659-0.851] | 1.0454 [0.971-1.125] | 0.2383 |
| Neutrophils | 135 | 23 | 0.0266 | 5 | 0.72 [0.616-0.824] | 0.9923 [0.905-1.089] | 0.8706 |
| Lymphocytes | 135 | 23 | - | - | - | - | - |
| **Prediction of intubation and ventilation** |  |  |  |  |  |  |  |
| MR-proADM | 135 | 16 | 22.648 | 5 | 0.793 [0.687-0.899] | 2.2457 [1.609-3.134] | 1.94e-6 |
| CRP<sup>1</sup> | 135 | 16 | 18.318 | 5 | 0.803 [0.713-0.893] | 1.0088 [1.005-1.013] | 1.87e-5 |
| PCT<sup>2</sup> | 116 | 16 | 2.241 | 5 | 0.715 [0.617-0.813] | 1.0558 [0.983-1.134] | 0.134 |
| Neutrophils | 135 | 16 | 0.4543 | 5 | 0.665 [0.563-0.767] | 1.0254 [0.953-1.103] | 0.500 |
| Lymphocytes | 135 | 16 | 0.2663 | 5 | 0.67 [0.554-0.786] | 0.8260 [0.400-1.707] | 0.606 |

<sup>1</sup> C-reactive protein
<sup>2</sup> Procalcitonin

Although PCT was significant for univariate Cox regression analysis (Wald 11·54, HR 1·0898 [1·037-1·145]), the assumption of proportional hazards was not supported for PCT. Rather than attempting to estimate a time-dependent coefficient in order to include covariates with time-varying coefficients multivariate analysis was not performed. Both MR-proADM (Wald 23·735, HR 1·809[1·454-2·407]) and CRP (Wald 12·166, HR 1·0069[1·003-1·011]) remained significant for predicting 30 day mortality following adjustment for age, cardiovascular disease, renal disease and neurological disease (Table 3). Similarly, lymphocyte count was significant in predicting 30 day mortality after adjusting for co-variates (Wald 6·6564, HR 1·1213[1·028-1·223]).

**Table 3.** Univariate area under receiver operating characteristic curve data for predicting various outcomes from baseline bloods

| Biomarker | Patients (N) | Cases (N) | D.F. | AUC [95% CI] |
| --- | --- | --- | --- | --- |
| 30 day mortality |  |  |  |  |
| MR-proADM | 135 | 30 | 1 | 0.8441 [0.7761-0.9122] |
| CRP<sup>1</sup> | 135 | 30 | 1 | 0.6132 [0.5066-0.7198] |
| PCT<sup>2</sup> | 116 | 26 | 1 | 0.6212 [0.4963-0.7460] |
| Neutrophils | 135 | 30 | 1 | 0.5503 [0.4356-0.6650] |
| Lymphocytes | 135 | 30 | 1 | 0.5629 [0.4373-0.6884] |
| Critical Care Admission |  |  |  |  |
| MR-proADM | 135 | 31 | 1 | 0.594 [0.4873-0.7007] |
| CRP<sup>1</sup> | 135 | 31 | 1 | 0.7238 [0.6213-0.8263] |
| PCT<sup>2</sup> | 116 | 31 | 1 | 0.6856 [0.5771-0.7941] |
| Neutrophils | 135 | 31 | 1 | 0.6158 [0.5104-0.7213] |
| Lymphocytes | 135 | 31 | 1 | 0.5296 [0.4147-0.6446] |
| NIV |  |  |  |  |
| MR-proADM | 135 | 23 | 1 | 0.7189 [0.617-0.8209] |
| CRP<sup>1</sup> | 135 | 23 | 1 | 0.722 [0.6068-0.8373] |
| PCT<sup>2</sup> | 116 | 22 | 1 | 0.7706 [0.6734-0.8677] |
| Neutrophils | 135 | 23 | 1 | 0.5615 [0.4505-0.6726] |
| Lymphocytes | 135 | 23 | 1 | 0.5646 [0.4476-0.6817] |
| Intubation and Ventilation |  |  |  |  |
| MR-proADM | 135 | 16 | 1 | 0.75 [0.6434-0.8566] |
| CRP<sup>1</sup> | 135 | 16 | 1 | 0.7545 [0.6059-0.903] |
| PCT<sup>2</sup> | 116 | 16 | 1 | 0.8497 [0.7565-0.9429] |
| Neutrophils | 135 | 16 | 1 | 0.6476 [0.5104-0.7848] |
| Lymphocytes | 135 | 16 | 1 | 0.468 [0.324-0.612] |

**Table 4.** Multivariate area under receiver operating characteristic data for predicting various outcomes from baseline bloods

| Biomarker | Patients (N) | Cases (N) | D.F. | AUC [95% CI] | Bootstrap |
| --- | --- | --- | --- | --- | --- |
| 30 day mortality |  |  |  |  |  |
| MR-proADM | 135 | 30 | 5 | 0.874 [0.8085-0.9395] | 0.01727 |
| CRP<sup>1</sup> | 135 | 30 | 5 | 0.8441 [0.769-0.9192] | 0.1464 |
| PCT<sup>2</sup> | 116 | 26 | 5 | 0.8406 [0.7619-0.9193] | 0.06908 |
| Neutrophils | 135 | 30 | 5 | 0.8394 [0.7667-0.9121] | 0.1417 |
| Lymphocytes | 135 | 30 | 5 | 0.8448 [0.7716-0.918] | 0.1056 |
| Critical Care Admission |  |  |  |  |  |
| MR-proADM | 135 | 31 | 5 | 0.7466 [0.6553-0.8379] | 0.1457 |
| CRP<sup>1</sup> | 135 | 31 | 5 | 0.8026 [0.7256-0.8796] | 0.012 |
| PCT<sup>2</sup> | 116 | 31 | 5 | 0.7632 [0.6747-0.8517] | 0.1465 |
| Neutrophils | 135 | 31 | 5 | 0.7252 [0.6359-0.8145] | 0.2458 |
| Lymphocytes | 135 | 31 | 5 | 0.7317 [0.6413-0.8221] | 0.2132 |
| NIV |  |  |  |  |  |
| MR-proADM | 135 | 23 | 5 | 0.8544 [0.7727-0.9362] | 0.000375 |
| CRP<sup>1</sup> | 135 | 23 | 5 | 0.8139 [0.7374-0.8904] | 0.001052 |
| PCT<sup>2</sup> | 116 | 22 | 5 | 0.7872 [0.6884-0.8861] | 0.03331 |
| Neutrophils | 135 | 23 | 5 | 0.7418 [0.6341-0.8496] | 0.07907 |
| Lymphocytes | 135 | 23 | 5 | 0.7519 [0.6461-0.8578] | 0.05353 |
| Intubation and Ventilation |  |  |  |  |  |
| MR-proADM | 135 | 16 | 5 | 0.812 [0.7071-0.9169] | 0.005858 |
| CRP<sup>1</sup> | 135 | 16 | 5 | 0.8183 [0.7176-0.9189] | 0.002934 |
| PCT<sup>2</sup> | 116 | 16 | 5 | 0.7256 [0.6193-0.8319] | 0.1324 |
| Neutrophils | 135 | 16 | 5 | 0.6817 [0.576-0.7874] | 0.2557 |
| Lymphocytes | 135 | 16 | 5 | 0.6838 [0.5664-0.8012] | 0.2675 |

MR-proADM offered a significant improvement in predicting 30 day mortality over a model combining age, renal disease, cardiovascular disease and neurological disease with an AUC of 0·874 (p=0·01727), whereas CRP did not offer an improvement (p=0·1464).

### Intubation and Ventilation

Elevated MR-proADM (Wald 12·37, HR 1·6015[1·232-2·082]) and CRP (Wald 16·57, HR 1·0069 [1·004-1·010]) were both associated with an increase in the need for intubation and ventilation when performing univariate cox regression. No firm conclusions are offered here from a multivariate analysis for prediction of need for intubation and ventilation due to the lower number of events per predictor variable, though the results are suggestive for both MR-proADM and CRP having efficacy in predicting the need for intubation.

### Admission to Critical Care

On univariate analysis only CRP demonstrated a significant association with a requirement for admission to critical care (Wald 15·83, HR [1·0055-1·008]), which was maintained once controlled for covariates, however proportionality was affected by renal disease (Chisq 4·2243, p=0·04), which was therefore taken out by stratification (Wald 22·062, HR 1·0078[1·005-1·011]). Although insignificant on univariate analysis MR-proADM (Wald 3·2, HR 1·257[0·978-1·615]) did exhibit a significant association with a requirement for critical care once covariates were controlled for (Wald 10·151, HR 1·5451 [1·182-2·019]).

Only CRP provided an improvement over the previously described combined model with an AUC of 0·8026 (p=0·012).

### Non-Invasive Ventilation

Once again elevated CRP (Wald 15·53, HR 1·0061[1·003-1·009]) and MR-proADM (Wald 11·62, HR 1·4992[1·188-1·892]) were associated with requirement for NIV following univariate analysis, with both CRP (Wald 18·293 HR 1·0081[1·004-1·012]) and MR-proADM (Wald 30·393, HR 2·5230[1·816-3·506]) both retaining significance on multivariate analysis. Lymphocyte count was not a constant hazard and so further analysis was omitted rather than estimating a time-dependent coefficient.

Both CRP and MR-proADM offered an improvement over the combined model with AUCs of 0·8544 (p=0·000375) and 0·8139 (p=0·001052) respectively.

## Discussion

Our study demonstrates that use of biomarkers, both established and novel, has significant potential to support clinical decisions and aid in identification of COVID-19 patients at both high and low risk for disease progression. Of the biomarkers evaluated, elevated MR-proADM and CRP concentrations showed the greatest potential for clinical application for utilisation to aid prediction of 30-day mortality, critical care admission requirement and need for both invasive and non-invasive ventilation.

Of these, on multivariate regression analysis MR-proADM concentration showed the strongest association with an increased 30-day mortality, requirement for non-invasive and invasive ventilation, with CRP showing the strongest association with critical care admission. Our data are also suggestive of an association between elevated MR-proADM and CRP concentration with a progression to being intubated and ventilated, though more cases are required to verify this by controlling for covariates. This is consistent with previous studies that have shown increased MR-proADM levels to be predictive of intensive care requirement in septic patients.^17^ Gonzalez del Castillo *et al*. identified MR-proADM to have the strongest association for ICU admission and mortality compared to other blood biomarkers in bacterial infections; with concentrations of >1·77nmol/L having significantly higher rates of ICU stay (8·1% vs 1·6%; p<0·001) and disease progression (29·7% vs 4·9%; p<0·001) compared to low MR-proADM concentrations.^18^ Application of these and our own findings, applying previously derived thresholds of 0·88 and 1·54nmol/L may support clinicians with decisions regarding earlier transfer of selected patients to wards with staff with experience in non-invasive ventilation, earlier transfer to critical care and dependent on emerging trial data, potentially those who are at low risk of deterioration suitable for ambulant monitoring. Similarly, subject to future studies, biomarker concentrations may have significant value in signalling patients suitable for earlier commencement of targeted therapies, including antiviral treatments (e.g. remdesivir) or immunomodulation with high dose steroids or interleukin-6 inhibitors (e.g. tocilizumab & sarilumab); in the NHS the latter are subject in the UK to specific inclusion criteria which includes physiological derangement and requirement for respiratory support.^19,20^

Previous studies examining the clinical utility of MR-proADM have focussed on predicting adverse events, primarily in patients with a diagnosis of community-acquired pneumonia and elevated MR-proADM levels,^4^ reflecting the development of organ dysfunction. However, these patients typically have had low rates of severe viral infection.^11, 21^ We have shown commensurate results in individuals diagnosed with SARS-CoV-2 infections. This may be explained by the development of endothelial dysfunction during SARS-CoV-2 infection, for which endoreticular stress and the activation of the unfolded protein response is a significant inducer. Endothelial dysfunction is associated with complications of Covid-19 including ARDS, thromboembolism and vascular disease, with Adrenomedullin known to be a regulator of vascular tone and the integrity and stability of the endothelial barrier.

The early recognition of potential for deterioration in patients infected with SARS-CoV-2 is invariably challenging given the varied clinical course of the disease; particularly with the currently unpredictable potential for deterioration during the second and third weeks of disease. As such, a reliable indicator for early identification of patients at risk of deterioration or a more severe illness trajectory could be of great value, in allowing for earlier targeted intervention or escalation of treatment.

The study has several limitations. Primarily, a higher event per variable ratio is desirable, fortunately however stratification was only needed for one covariate. Greater power could be achieved through higher participant numbers or through examining serial results over time. A multi-centre study with more patients could also allow for prediction of progression from NIV to intubation. It is a retrospective study; it remains to be seen whether application of such biomarkers into clinical diagnostic and management pathways will deliver the potential expected benefits. Clinician confidence in use of such biomarkers to support decisions has to be developed and evidenced in clinical practice.

MR-proADM and CRP may have other clinical utility worth exploring, such as whether measurement of these has use in facilitating earlier discharges^11^ in individuals with low concentrations or whether a delta change in concentrations could be used to guide decisions on weaning ventilatory support and also the de-escalation or stopping of antibiotics to mitigate against potential future antimicrobial resistance.

## Conclusions

Biomarker measurement, in particular MR-proADM, in patients infected with SARS-CoV-2 upon presentation to secondary care shows significant promise in allowing early identification of those at increased risk of disease progression and mortality, possibly providing a tool to support clinical decisions regarding therapeutic interventions and level of care setting. Additionally, adopting use of biomarkers such as MR-proADM in the acute setting carries potential in allowing identification of those at low-risk for severe disease, facilitating discharge decisions. Further prospective, multi-centre studies are needed to investigate possible exciting clinical applications of this and other biomarkers.

### Key Messages

- Admission MR-proADM predicts 30-day mortality in SARS-CoV-2 infection
- Admission MR-proADM predicts critical care requirement in SARS-CoV-2 infection
- Admission MR-proADM predicts NIV requirement in SARS-CoV-2 infection
- Admission CRP predicts 30-day mortality, NIV and critical care requirement
- Admission PCT and White Cell Count do not predict clinical outcomes

## Data Availability

All data used is available upon request to corresponding author

## Author Contributions

NAM, MY, KG and NC contributed to the study conception and design. Material preparation was performed by TL. Data collection was performed by RW, MM, BB, GV and JL. Analysis was performed by NAM and PP. The first draft of the manuscript was written by NAM. RW, SPK, CT, VGA, KS, KG and NC commented on previous versions of the manuscript. All authors read and approved the final manuscript.

## Ethical approval

Local Hampshire Hospitals NHS Foundation Trust R&D approval was obtained. HRA Approval was sought who deemed an ethical review was not necessary. HRA Approval was subsequently granted for this project (IRAS project ID: 299130). Anonymised confidential patient information was used under the COVID-19 COPI notice issued by the UK Department of Health and Social Care

